# Shared and Distinct Brain Regions in Anxiety, Depression, and Neuroticism: A Multi-Phenotypic Approach Using Data from the UK Biobank

**DOI:** 10.1101/2025.08.26.25334447

**Authors:** Chelsea Sawyers, Sarah E. Benstock, Brad Verhulst, Lisa K. Straub, John M. Hettema

## Abstract

**Background:** Anxiety and depression are highly comorbid and share common clinical symptoms and etiological factors, particularly through overlapping genetic and environmental risk. Neuroticism strongly correlates with both anxiety and depression, and genetic studies suggest it accounts for a significant portion of the shared genetic risk between these disorders.

**Methods:** This study used data from 46,541 participants from the UK Biobank with available imaging data including 4,355 with a lifetime anxiety disorder and 8,559 with major depressive disorder to investigate the neuroanatomical correlates of anxiety, depression, and neuroticism with a focus on identifying shared and distinct associated brain regions across these phenotypes. Using structural MRI data, we examined cortical thickness (CT), surface area (SA), and subcortical volume associations with both subscale-specific and full measures of neuroticism and these disorders.

**Results:** Findings revealed significant associations between anxiety disorders and thinner CT in the left insula, posterior cingulate, and several frontal and temporal regions. Depression was associated with increased right caudate volume, decreased SA in the pericalcarine and cuneus, and thinner lateral occipital and posterior cingulate CT. Neuroticism and its subscales showed more widespread brain involvement, with significant associations in areas including the bilateral caudate, medial orbitofrontal cortex, and precentral gyrus. Notably, neuroticism measures demonstrated stronger associations than diagnosis-specific outcomes, emphasizing the utility of dimensional approaches. Only one significant association (thinner left posterior cingulate) was shared between anxiety and depression, but several trends were observed.

**Conclusions:** Our findings further support a partial neurobiological sharing between anxiety and depression with neuroticism providing a nuanced framework for their connections.

## INTRODUCTION

Anxiety (ANX) and depressive disorders are highly prevalent, persistent, and cause extensive disability worldwide(1). They are highly comorbid with each other over an individual’s lifetime such that more than half of those with an ANX eventually develop a depressive disorder and vice versa (2–4). Such comorbidity complicates and exacerbates their detrimental effects on one’s life by causing worsening distress, impairment, and poorer treatment response. ANX and depression are strongly related to each other in areas of overlapping clinical symptoms, etiologic factors, and treatment protocols. In particular, they share both genetic and environmental risk factors(5,6). Interestingly, the normal personality trait of neuroticism predicts anxiety-depression comorbidity(7,8). That is not surprising, since neuroticism predicts an individual’s negative emotionality at baseline and in reaction to stress(9). Genetic studies suggest that a substantial portion of the shared genetic risk between ANX and depressive disorders is attributable to genetic variation in neuroticism(10–12). In other words, there are genetic factors common to neuroticism, ANX, and depressive disorders that help explain their covariation. Furthermore, the classic formulation of neuroticism by Eysenck includes subsets of items that are more specific to anxious worry in response to stress and others more specific to depressive affect. Genome-wide association studies have begun to study these distinct but related aspects of neuroticism(13).

In addition to genetics, neuroimaging studies support a shared biological basis between ANX and depressive disorders with overlapping regional differences in brain structure(14,15) or function(14,16) that are involved in both types of disorders. With inconsistent findings across primary studies, meta-analyses for anxiety have struggled to detect converging results in structural(17) and task activation studies(18,19). However, depression has shown more consistent structural findings(20,21), and resting-state connectivity appears to fare even better(22,23). This is most likely due in part to differing sample sizes of the original studies(24). The most consistent findings shared between these disorders include differences in cortical thickness in the orbital frontal cortex (OFC), posterior and anterior cingulate, and insula cortices (21,23) and in the amygdala and hippocampus subcortical volumes(20,23). A few neuroimaging studies have focused on brain differences that covary with neuroticism, finding variations in pars opercularis, left frontal and parietal cortices(25), and anterior cingulate and medial prefrontal cortices(26). Some of these differences overlap with regions reported for ANX(23), depressive disorders(21), or both. However, only a single study systematically examined the shared and specific brain regional differences among neuroticism, ANX, and depression in a relatively small sample of young adults with subsyndromal to syndromal mood and anxiety symptoms(25). No imaging studies have analyzed the effects of the neuroticism subscales, anxious worry and depressed affect, on brain structure.

In this study, we sought to answer the following questions. (1) What brain regions of interest (ROIs) are associated with neuroticism, ANX, and depression? (2) Which ROIs are shared and which are specific to each of these phenotypes? (3) Do the neuroticism subscales help explain the pattern of overlap? We used the phenotypic and neuroimaging data available from the UK Biobank study to maximize power and minimize heterogeneity.

## MATERIALS AND METHODS

### Participants

The current study used data from the UK Biobank (27) (application 57923) accessed on November 15^th^ 2023. The UK Biobank is a large, phenotypically detailed dataset including individuals aged 40 to 70 years old. Participants were included in the current study if they had both data from the Mental Health Questionnaire (28) and structural MRI data that had been processed with FreeSurfer (29). A total of 46,541 participants had imaging data that passed the standard quality controls of UK Biobank and information about their mental health status.

### Psychiatric Phenotypes

We examined a series of psychiatric phenotypes defined from the UK Biobank: neuroticism, depressed affect in neuroticism, anxious worry in neuroticism, any lifetime anxiety disorder, and lifetime major depression - see Supplement for details. Neuroticism was defined by a pre-calculated summed score using 12 dichotomous questions from the revised short form of the Eysenck Personality Questionnaire (30). We subsequently divided the 12 items used to calculate the total summed score into a depressive affect score (DepAff) and an anxious worry score (Worry) previously defined (31).

In addition to these dimensional phenotypes, we created case-control cohorts for the two psychiatric outcomes. The broad any lifetime anxiety disorders (ANX) group was determined using three sources of information available. Individuals were considered cases if they (1) self-reported a professional diagnosis of a specific anxiety disorder (panic disorder, social anxiety disorder, agoraphobia, or specific phobia), (2) received an ICD-9 or ICD-10 diagnosis for an anxiety disorder, or (3) met Composite International Diagnostic Interview short-form (CIDI-SF) diagnostic criteria (32,33) for generalized anxiety disorder (GAD). ANX controls did not meet any of the criteria for being a case or answered “no” to the CIDI-SF screening questions for GAD.

The second case-control outcome was for lifetime major depressive disorder (MDD). Individuals were considered cases if they met CIDI-SF diagnostic criteria for MDD or received an ICD-9 or ICD-10 diagnosis for MDD. Controls for the MDD phenotype were constructed similarly to the controls for the ANX phenotype, where they did not meet diagnostic criteria or answered “no” to the CIDI-SF screening questions for MDD. Additionally, individuals who self-reported a professional diagnosis of schizophrenia, bipolar disorder, autism spectrum disorder, or selected that they preferred not to answer were removed prior to all analyses.

### Imaging Data

We included structural imaging data processed with FreeSurfer and collected between 2014 and 2018. A comprehensive description of the imaging data collection and processing procedures can be found elsewhere(34,35). The cortical thickness (CT), cortical surface area (SA), and subcortical segmentations of the Desikan-Killiany-Tourville atlas(36) were used in these analyses. Given that SA and CT show distinct independent genetic influences both globally and locally(37–39), we opted to examine SA and CT measures separately rather than utilizing cortical gray matter volumes which appear to be dominated by SA more so than CT(39).

### Statistical Analysis

All statistical analyses were conducted in R (40). We applied linear regression using the outcome phenotypes as the main predictor for each ROI with sex, age, age^2^, the interaction of sex and age, the interaction of sex and age^2^, and total intracranial volume as covariates. Age^2^ and its interaction with sex were included to account for any non-linear influences of age on the ROIs. Due to the narrow age range of the sample (40 to 70 years), we recentered the sample based on the mean age. Post-hoc corrections for testing 140 ROIs were conducted using a Bonferroni-adjusted p-value (0.05/140; p < 0.00036)(41).

## RESULTS

Sample characteristics of the data available for analysis are provided in Table 1. We ran a series of analyses examining the association of CT and SA of cortical ROIs and volume of subcortical ROIs across the five primary phenotypes. Table 2 lists the regional associations by phenotype after controlling for model covariates. Effect sizes are displayed as t-scores for differences and their p-values. Significant associations after adjusting for multiple testing are indicated in bold (P_adj_ = 0.00036). Measures (subcortical volume or SA or CT) are included in the table if they show significant association for at least one outcome phenotype. The columns are arranged from left to right from ANX through neuroticism dimensions to MDD. Figures 1 and 2 display these associations projected onto the associated brain region space. Figure 1 shows the effect size for cortical SA or CT ROIs across the phenotypes in panels A (ANX), B (Worry), C (Neuroticism), D (DepAff), and E (MDD). Figure 2 does the same for associated subcortical volumes in panels A (MDD), B (DepAff), and C (Neuroticism); ANX and Worry did not show associations with subcortical volumes.

**Figure 1.**
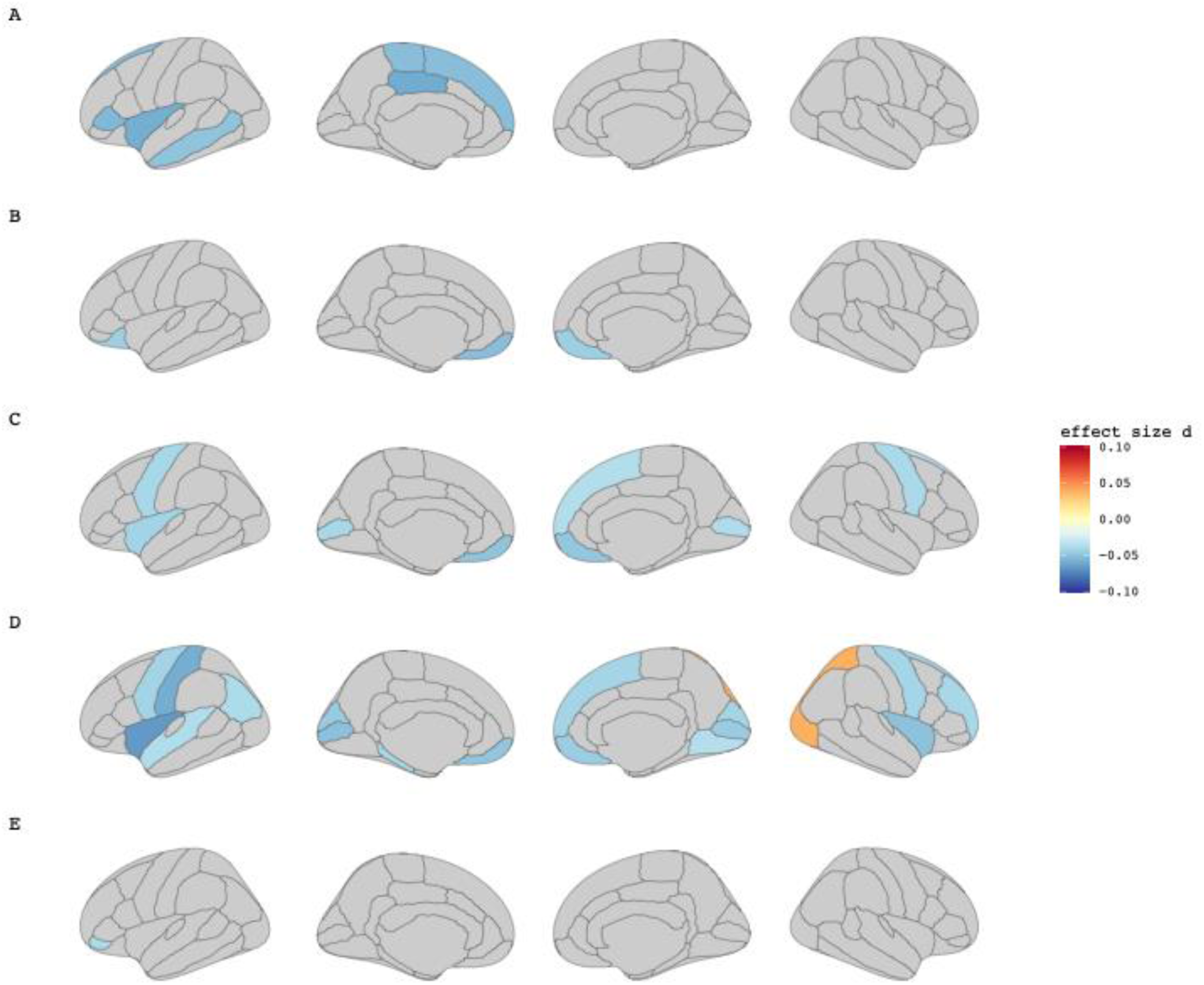
Cohen’s d Effect Size for Regions with Significant Cortical Thickness or Surface Area Across Anxiety, Depression and Neuroticism. Each panel illustrates the effect size of the phenotype for each region regions with significant cortical thickness or surface area across A) Anxiety, B) Worry, C) Neuroticism, D) Depressive Affect, and E) Depression (MDD). Effect sizes were corrected for age, sex, and intracranial volume.

**Figure 2.**
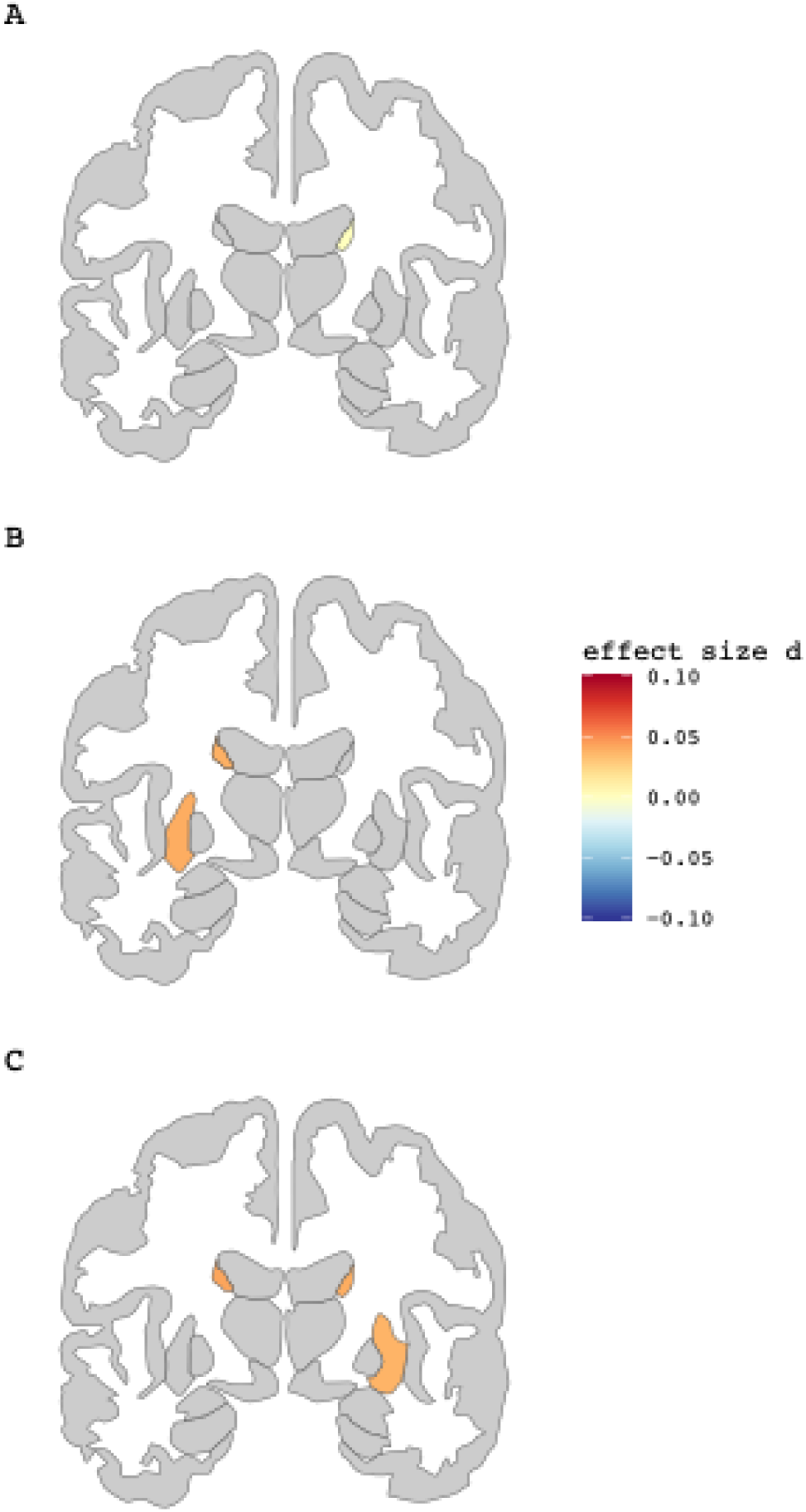
Cohen’s d Effect Size for Subcortical Regions with Significant Results. Each panel illustrates the effect size of the phenotype for each region with significant association to A) Depression (MDD), B) Depressive Affect (DepAff) and C) Neuroticism. Effect sizes were corrected for age, sex, and intracranial volume.

Neuroticism and its subscales showed notably more associations than either of the clinical phenotypes. In particular, we saw the greatest number of significant associations for DepAff (23 in total). These include (1) larger volumes of caudate (bilateral; B) and putamen (B), (2) smaller cortical areas of pericalcarine (B), cuneus (B), postcentral (left; L), inferior parietal (B), superior frontal (right; R), rostral middle frontal (R), and lingual (R), (3) thinner cortex in the insulae (B), parahippocampal (L), superior temporal (L) regions, and (4) thicker cortex in the lateral occipital (L) and superior parietal (R) regions. Several of these overlap with the same findings for MDD, while only one (thinner left insula) overlaps with the same finding in ANX. In total, there were ten ROIs with overlap between these associations for DepAff and the full neuroticism score. Figure 3 displays the patterns of overlap using an upset plot. With far fewer findings, Worry was significantly associated with a smaller area of medial OFC bilaterally; this was consistent across all three neuroticism phenotypes, with a trend for ANX. Worry also showed an isolated association with thinner lateral OFC (left only).

**Figure 3.**
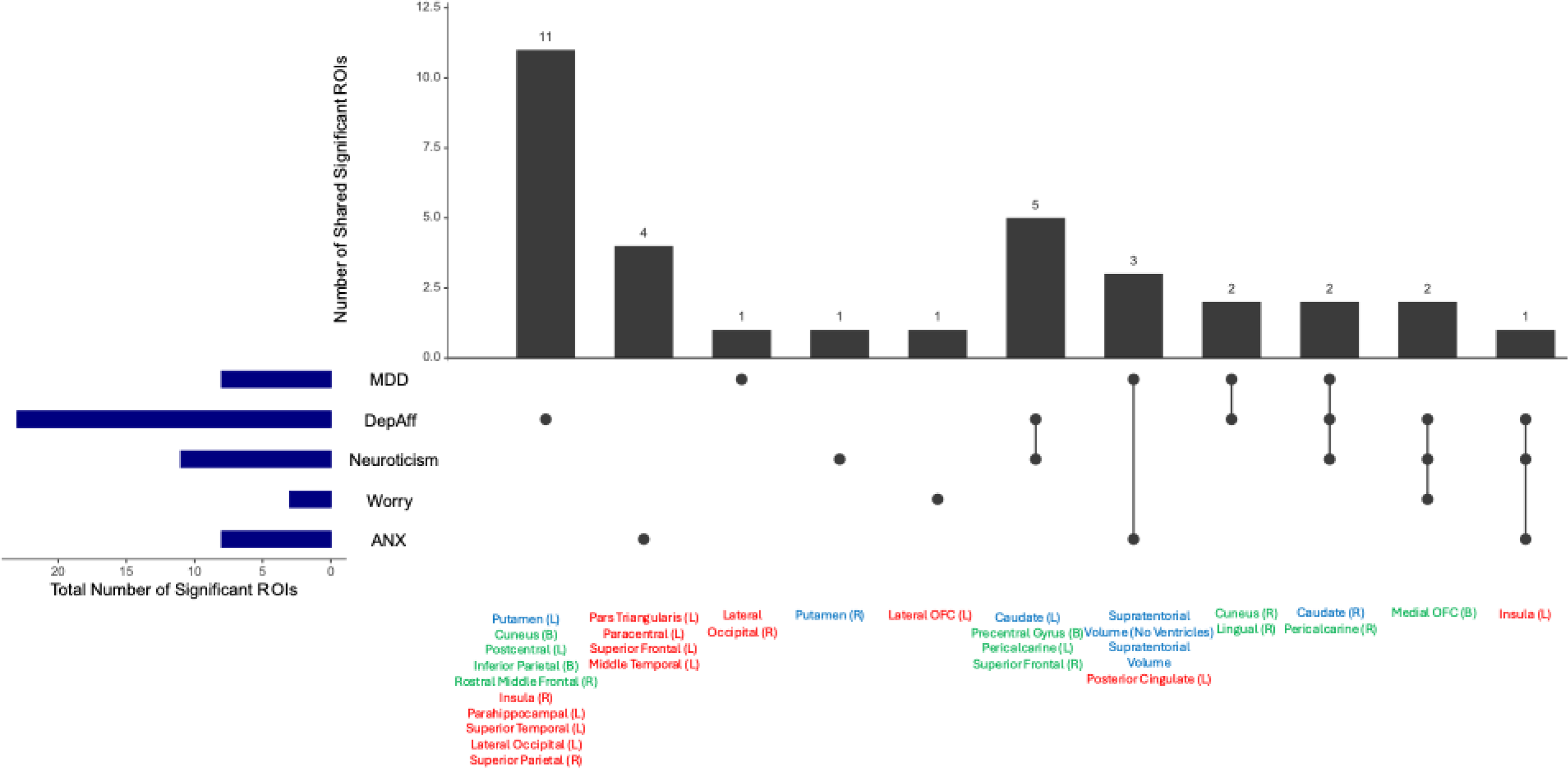
Upset Plot for Significant ROIs for each Outcome. The dark blue bars on the bottom left indicates how many total ROIs are significant for each outcome. The right side of the plot indicates the number of overlapping, significant ROIs for the outcomes. The significant ROIs are also listed before each point and corresponding bar, where blue represents subcortical volumes, green represents cortical area, and red represents cortical thickness.

Turning to the clinical disorders, MDD showed significant associations with smaller SA of the right pericalcarine, cuneus, and lingual gyri, thinner left posterior cingulate and thicker right lateral occipital gyrus, a larger right caudate volume and an overall smaller supratentorial volume. ANX was significantly associated with an overall smaller supratentorial volume and thinner left-sided insula, pars triangularis, paracentral, superior frontal, middle temporal and posterior cingulate cortices, the last being the only association shared with MDD.

## DISCUSSION

Given the strong clinical and etiological overlap between ANX, MDD, and neuroticism, we aimed to identify associated brain region differences and to determine whether these regions are specific to, or shared between, these phenotypes. Additionally, we explored whether subscales within neuroticism would explain any observed pattern of overlap. We found that thinner cortical thicknesses in the left insula, posterior cingulate, pars triangularis, paracentral gyrus, superior frontal gyrus, and middle temporal gyrus were all associated with ANX, whereas MDD was associated with larger right caudate volume and a smaller SA of the right pericalcarine, cuneus, and lingual gyri, thinner left posterior cingulate and thicker right lateral occipital gyrus. Neuroticism and its subscales were associated with bilateral caudate volume, right putamen volume, bilateral area of the precentral gyrus, medial OFC, pericalcarine, cuneus, inferior parietal, left area of the postcentral gyrus, and right area of the superior frontal, rostral middle frontal, and lingual gyri, as well as cortical thickness of the bilateral insula, left lateral OFC, lateral occipital, superior temporal, parahippocampal gyri, and the right superior parietal gyrus.

Our findings showed that the ROIs significantly associated with ANX were predominantly in the frontal lobe, with the insula and a portion of the cingulate cortex also reaching significance. A previous meta-analysis examining GAD specifically by the ENIGMA-Anxiety Working Group found no significant CT or SA ROI associations(17); smaller, individual studies show inconsistent findings regarding CT of the cingulate(42,43), orbitofrontal(42,44,45) and temporal(46) regions. Our results diverge from a previous meta-analysis of specific phobias(47) with no overlap between the results of the two studies. Due to their lack of direct assessment and low rates of clinical presentation, phobias were not well represented in UK Biobank. A meta-analysis examining anxiety severity and voxel-based morphometry (VBM) measures in GAD(48) found reduced gray matter (GM) volume in the right insula, whereas two other meta-analyses examining GM volumes with any anxiety diagnosis(49,50) both found bilateral reduction of GM volume in the insula. While the direction of effect in the GAD-specific meta-analysis is similar, our findings show an opposite lateralization pattern. An older VBM meta-analysis found reduced GM volume in GAD within the pars triangularis and middle temporal gyrus(23), consistent with our findings of decreased CT in these areas; however, those findings were not replicated in the larger meta-analysis(48). Although findings of structural differences in the posterior cingulate cortex (PCC) between anxiety and controls have been limited, task-based MRI analyses have demonstrated that deactivation of the PCC may be involved in cognitive deficits associated with clinical anxiety(51–53).

Our results demonstrated an interesting pattern where left hemisphere ROIs accounted for all six of the significant ROI associations with ANX, whereas only one of the MDD associated ROIs was in the left hemisphere. Across all the phenotypes, medial and more caudal ROIs predominantly showed bilateral significance (8 of 12 ROIs), whereas significant ROIs within the frontal, temporal, and insular lobes demonstrated a left-sided lateralization (8 of 9 ROIs). Electrophysiological studies suggest this disruption of brain lateralization can be found across psychiatric disorders(54,55), whereby worry (anxious apprehension)(56–59) is predominantly associated with left-sided activity in the frontal lobe. The same pattern can be found in rodent models, which demonstrate hemispheric asymmetry in fear conditioning and anxiety-inducing rat(60) paradigms. Two main theoretical approaches to this frontal brain lateralization have emerged over time: a worry/avoidance model and an emotional valence (positive/negative) model(61). Our findings possibly support the worry/avoidance model, where worry cues lateralization of the frontal brain activity leftward. Additionally, our findings of only the ANX group demonstrating the left frontal lateralization pattern is consistent with previous work that linked GAD specifically with greater left frontal activity(57).

Overall, our examination of MDD with cortical and subcortical structures replicated several findings from previous meta-analyses(21,62). Our finding of thinner posterior cingulate cortex is consistent with meta-analyses in adults, while the smaller SA of the right pericalcarine and right lingual gyrus aligns with previous work in adolescents(21). We found a smaller SA was only significant in the right cuneus (although trending in the left) compared to previous meta-analysis in adolescents that found only the left was significant. The DepAff subscale in our study is consistent with that left-hemisphere-specific cuneus finding. Also of note, the DepAff subscale aligns with previous meta-analytic findings from the ENIGMA Major Depressive Disorder Working Group of decreased thickness in the insula, temporal, OFC, and posterior cingulate cortices in MDD(21).

We found a common pattern of association for neuroticism and MDD in the ventral striatum, where bilateral caudate volumes exhibited the strongest association with neuroticism, remained significant for DepAff, and were only significant for MDD in the right caudate with left and right caudate trending for ANX after correcting for multiple tests. Previous meta-analyses of depression had found no association with caudate volume; however, meta-analyses of task-based functional MRI have shown alternatively reported a reduction in activation of the ventral striatum in depressed patients and greater activation depending on the sub-region(63). Finding associations within the ventral striatum and depression in our study possibly aligns with the common clinical presentation of anhedonia (reduced positive affect) as well as with VS activation patterns observed in depressed patients(64–66). The significance pattern observed in the caudate might be consistent with functional genomic analyses that showed a polygenic score for neuroticism moderated caudate activation during anticipation of a reward(67). Resting-state functional connectivity analyses also show that high neuroticism resulted in greater activation and lower arousal thresholds for the right caudate(68).

Another pattern of association emerged for neuroticism and Worry with bilateral surface area mOFC, with ANX and MDD trending in the same direction but not surviving multiple testing corrections. This is consistent with a previous study that found the same direction of effect for neuroticism and mOFC volume in adolescent females(69).

Overall, Table 2 indicates 15 regions in total with significant associations across more than one of the five phenotypes; not surprisingly, those are dominated by sharing across the neuroticism groups. However, we note that for each row in Table 2, there is a consistent trend across many of the phenotypes. For example, the caudate volume is trending larger in all these phenotypes although only reaching significance for neuroticism, DepAff, and MDD. Additionally, the direction of effect is consistent across ANX, neuroticism and DepAff for the left insular and left posterior cingulate thickness. Figure 3 depicts these significant areas of phenotypic overlap with an Upset Plot.

The neuroticism measures showing more associations overall compared to the case-control analyses also highlights the important role of increased power when using continuous measures compared to binary outcomes. Not only are more individuals available for analyses in this sample, but the information each participant brings is greater for continuous variables. This provided additional power to detect differences beyond the case/control ANX and MDD analyses in this already adequately-powered sample.

Previous work examining neuroanatomical profiles of depression and anxiety also suggests a shared etiology underlying the common neurotic symptom dimensions of anxiety and depression(15). Transdiagnostic meta-analyses(70,71) across a broad range of psychiatric diagnoses indicate a shared pathology across neuropsychiatric diseases broadly with limited weak neurobiological ‘signal’ for specific disorders. Our findings are consistent with this trend in that, although a thinner PCC was significant for both ANX and MDD, the broader neuroticism measures showed more associations overall, and more overlap was observed between neuroticism and each individual clinical phenotype than between the disorders themselves.

These findings should be interpreted in the context of several limitations. First, the age of participants (40-70 years) is a limited portion of the lifespan. These results may differ if examined in a pediatric study when the brain is undergoing substantial developmental changes or in a sample of young adults closer to the peak age of onset for anxiety and depression.

However, the older sample and expected age-at-onset for these disorders provides reassurance that there should be limited conversion of control-to-case status. Second, only GAD and MDD were directly assessed by CIDI-SF, leaving identification of most of the anxiety disorders to ICD coding or self-report. This leaves open the possibility of missed-diagnoses in some individuals or potential misdiagnosis in others depending on the reporting reliability of each participant. Lastly, we did not account for medication use in analyses which could potentially influence the magnitude of associations found.

## CONCLUSIONS

In conclusion, our study sheds light on the complex neuroanatomical underpinnings of anxiety, depression, and neuroticism by identifying specific brain regions associated with each phenotype as well as shared regions across these conditions. This is the largest study to date to investigate brain regions associated with anxiety and the potential overlap with depression and neuroticism. While we observed distinct patterns, such as left hemisphere lateralization in anxiety and associations with the ventral striatum in depression; these phenotypes also demonstrated overlapping findings, particularly in areas like the caudate, insula, and PCC. Neuroticism revealed more widespread associations than the clinical phenotypes, supporting the value of using dimensional measures to capture nuanced brain-behavior relationships. Notably, our findings align with the view of a shared etiology between anxiety and depression reinforced by a partially shared neurobiological basis underlying these disorders. This work underscores the importance of studying transdiagnostic traits to better understand the underlying brain structures involved in psychiatric conditions.

## Data Availability

All data used in the present study are available from the UK Biobank.

https://www.ukbiobank.ac.uk/use-our-data/apply-for-access/

## Supplemental Materials

Supplementary information is available at Biological Psychiatry’s website.

## Acknowledgements

This research has been conducted using data from the UK Biobank, a major medical database (Application Number 57923). We would like to thank the participants who contributed their time and effort for their vital contribution to this research. This article was available via pre-print on MedRxiv. CS contributed to the conceptualization, methodology, writing-original draft, writing-review & editing, and visualization. SEB contributed to the methodology, formal analysis, writing-original draft, writing-review & editing, and visualization. BV contributed to the methodology, formal analysis, writing-review & editing. LKS contributed to the writing-original draft, and writing-review & editing. JMH contributed to the conceptualization, supervision, writing-original draft, and writing-review & editing.

