## Supplemental Tables for "Shared and Distinct Brain Regions in Anxiety, Depression, and Neuroticism: A Multi-Phenotypic Approach Using Data from the UK Biobank"

### All Neuroticism Items

| | |
|---|---|
| 1920-0.0 | Mood Swings |
| 1930-0.0 | Miserableness |
| 1940-0.0 | Irritability |
| 1950-0.0 | Hurt Feelings |
| 1960-0.0 | Feeling Fed Up |
| 1970-0.0 | Nervous |
| 1980-0.0 | Worrier |
| 1990-0.0 | Tense |
| 2000-0.0 | Embarrassed |
| 2010-0.0 | Suffer Nerves |
| 2020-0.0 | Loneliness |
| 2030-0.0 | Guilty |
| 20127-0.0 | Neuroticism summed score |

### Anxious Worry in Neuroticism Items

| | |
|---|---|
| 1970-0.0 | Nervous |
| 1980-0.0 | Worrier |
| 1990-0.0 | Tense |
| 2010-0.0 | Suffer Nerves |

### Depressed Affect in Neuroticism Items

| | |
|---|---|
| 1920-0.0 | Mood Swings |
| 1930-0.0 | Miserableness |
| 1960-0.0 | Feeling Fed Up |
| 2020-0.0 | Loneliness |

**Lifetime Major Depressive Disorder (MDD)**

*CIDI-SF MDD Items*

| | |
|---|---|
| 20446-0.0 | Depressed mood |
| 20441-0.0 | Anhedonia |
| 20536-0.0 | Weight change |
| 20532-0.0 | Sleep change |
| 20449-0.0 | Tiredness/low energy |
| 20450-0.0 | Worthlessness |
| 20435-0.0 | Difficulty concentrating |
| 20437-0.0 | Suicidal thoughts |
| 20436-0.0 | Fraction of day affected during worst episode of depression |
| 20439-0.0 | Frequency of depressed days during worst episode of depression |
| 20440-0.0 | Impact on normal roles during worst period of depression |
| 20448-0.0 | Professional informed about depression |
| 20546-0.1 | Substances taken for depression |
| 20547-0.1 | Activities undertaken to treat depression |

MDD Case: Participant endorsed 5 or more clinical symptoms including depressed mood (20446.0.0) and/or anhedonia (20441.0.0). Must also endorse mood persistence (20436.0.0 and 20439.0.0) and at least one form of distress & impairment (20440-0.0, 20448-0.0, 20546-0.1, 20547-0.1).

**OR**

Participant received an ICD-9 (311) or ICD-10 (F32.0-F32.9 and F33.0-F33.9) lifetime diagnosis of MDD from electronic health records

Supplementary Table 1. Cross-tabulation of participants who received an ICD-9 diagnosis vs. those who met our diagnostic criteria for MDD using CIDI-SF

| CIDI-SF \ ICD-9 | 0 | 1 |
|---|---|---|
| 0 | 135 | 4 |
| 1 | 252 | 16 |

Supplementary Table 2. Cross-tabulation of participants who received an ICD-10 diagnosis vs. those who met our diagnostic criteria for MDD using CIDI-SF

| CIDI-SF \ ICD-10 | 0 | 1 |
|---|---|---|
| 0 | 42,997 | 0 |
| 1 | 41,060 | 16,625 |

**Lifetime Any Anxiety Disorder**

*CIDI-SF Generalized Anxiety Disorder Items*

| | |
|---|---|
| 20421-0.0 | Ever felt worried, tense, or anxious for most of a month or longer |
| 20425-0.0 | Ever worried more than most people would in similar situation |
| 20420-0.0 | Longest period spent worried or anxious |
| 20417-0.0 | Tense, sore, or aching muscles during worst period of anxiety |
| 20419-0.0 | Difficulty concentrating during worst period of anxiety |
| 20422-0.0 | More irritable than usual during worst period of anxiety |
| 20423-0.0 | Keyed up or on edge during worst period of anxiety |
| 20426-0.0 | Restless during period of worst anxiety |
| 20427-0.0 | Frequent trouble falling or staying asleep during worst period of anxiety |
| 20429-0.0 | Easily tired during worst period of anxiety |
| 20537-0.0 | Frequency of difficulty controlling worry during worst period of anxiety |
| 20538-0.0 | Worried most days during period of worst anxiety |
| 20539-0.0 | Frequency of inability to stop worrying during worst period of anxiety |
| 20540-0.0 | Multiple worries during worst period of anxiety |
| 20541-0.0 | Difficulty stopping worrying during worst period of anxiety |
| 20542-0.0 | Stronger worrying (than other people) during period of worst anxiety |
| 20543-0.0 | Number of things worried about during worst period of anxiety |
| 20418-0.0 | Impact on normal roles during worst period of anxiety |
| 20428-0.0 | Professional informed about anxiety |
| 20549-0.0 | Substances Taken for anxiety |
| 20550-0.0 | Activities undertaken to treat anxiety |

ANX Case: Participant met CIDI-SF GAD criteria, meaning they endorsed having felt worried, tense, or anxious for most of a month or longer 20421-0.0 or 20425-0.0, worried most days during period of worst anxiety, and their longest period of anxiety was 6 or more months. Additionally, cases had 3 or more worry symptoms (20537-20543), at least 3 clinical associated symptoms (20417.0.0-20429.0.0 excluding 20420.0.0 and 20428.0.0), and at least 1 distress or impairment item (20418.0.0, 20428.0.0, 20549.0.0, 20550.0.0)

**OR**

Participant received an ICD-9 (F30.x) or ICD-10 (F40.0, F40.1, F40.2, F40.8, F40.9, F41.0, F41.1, F41.3, F41.8, F41.9) lifetime anxiety disorder diagnosis from electronic health records

**OR**

Participant self-reported a lifetime professional diagnosis of Social Anxiety, Agoraphobia, Panic Disorder, or Specific Phobia

Supplementary Table 3. Total Number of participants who self-reported a professional diagnosis (SRPD) of an anxiety disorder

| | |
|---|---|
| Social anxiety or Social phobia | 1,765 |
| Agoraphobia | 532 |
| Panic disorder | 8,426 |
| Specific phobia | 2,057 |
| Total with SRPD of an anxiety disorder | 11,043 |

Supplementary Table 4. Cross-tabulation of participants who received an ICD-9 diagnosis vs. those who self-reported a professional diagnosis of an anxiety disorder

| ICD-9 / Self-report | 0 | 1 |
|---|---|---|
| 0 | 2,283 | 43 |
| 1 | 52 | 0 |

Supplementary Table 5. Cross-tabulation of participants who received an ICD-10 diagnosis vs. those who self-reported a professional diagnosis of an anxiety disorder

| ICD-10 / Self-report | 0 | 1 |
|---|---|---|
| 0 | 477,740 | 10,398 |
| 1 | 10,300 | 743 |

Supplementary Table 6. Cross-tabulation of participants who self-reported a professional diagnosis of an anxiety disorder vs. those who met our CIDI-SF GAD criteria

| CIDI-SF \ Self-report | 0 | 1 |
|---|---|---|
| 0 | 92,050 | 0 |
| 1 | 20,744 | 11,043 |

Supplementary Table 7. Cross-tabulation of participants who received an ICD-9 diagnosis of an anxiety disorder vs. those who met our CIDI-SF GAD criteria

| CIDI-SF \ ICD-9 | 0 | 1 |
|---|---|---|
| 0 | 257 | 0 |
| 1 | 149 | 43 |

Supplementary Table 8. Cross-tabulation of participants who received an ICD-10 diagnosis of an anxiety disorder vs. those who met our CIDI-SF GAD criteria

| CIDI-SF \ ICD-10 | 0 | 1 |
|---|---|---|
| 0 | 92,050 | 0 |
| 1 | 20,646 | 11,141 |
